# A Comparative Analysis of Global Adult Tobacco Survey (GATS): Prevalence and Predictors of Tobacco Smoking in Seven Regions of India

**DOI:** 10.1101/2023.10.24.23297484

**Authors:** Vishal Tikhute

## Abstract

**Aim:** This article aims to assess the prevalence of tobacco smoking among the adult study population and also explore the association between socio-demographic factors and tobacco use in the six geographical regions of India.

**Methods:** This is a cross-sectional comparative research design that used nationally representative population-based household GATS-1 data on tobacco use in India. Six states representing the respective six geographic regions (one state per region) of India were selected using systematic random sampling. The prevalence of tobacco smoking among the study population was calculated and used for comparison between the six regions. Logistic regression was used to assess the association between socio-demographic factors and tobacco smoking in each zone.

**Results:** The North-eastern, Central, and Eastern regions had a high prevalence of tobacco smoking. Further, factors such as sex, place of residence, literacy, and working status had a significant association with current use of tobacco (smoking).

**Conclusion:** Despite there are policies and legal frameworks to control the use and sell of tobacco in India, the prevalence of tobacco smoking is high, particularly in the North-eastern, Central, and Eastern regions of India. Policies and community-based approaches to provide health education to reduce tobacco smoking are recommended.

---

Tobacco use is cause behind premature morbidity and mortality. ^1^ Tobacco kills more than 8 million people each year world-wide. ^1^ More than 8 millions of those deaths are the result of direct tobacco use while around 1.3 million are the result of non-smokers being exposed to second-hand smoke. ^1,2^ Cigarette smoking harms nearly every organ of the body, causes many diseases, and reduces the health of smokers in general. ^3^ According to WHO’s south East Asia region office, Tobacco use is a major risk factor for many chronic diseases including cancer, lung diseases, cardiovascular diseases and stroke. ^1,3^

Despite having greater contributor towards tobacco related morbidity and mortality; India has one of the highest tobacco users in the world both in number and relative share. ^4^ India is the second largest consumer and third largest producer of tobacco and a plethora of tobacco products are available at very low prices. ^5^ The country has a long history of tobacco use. Nearly 8–9 lakh people die every year in India due to diseases related to tobacco use. ^6^ Majority of the cardiovascular diseases, cancers and chronic lung diseases are directly attributable to tobacco consumption. ^3,6,7^ Almost 40 per cent of tuberculosis deaths in the country are associated with smoking. ^7^

## GLOBAL ADULT TOBACCO SURVEY (GATS) INDIA

The Global Adult Tobacco Survey – 2010, was carried out in all six geographical regions for both urban and rural areas of 29 states and two Union Territories (Chandigarh and Pondicherry) covering about 99.9 per cent of the total population of India. ^8^ The key findings from Global Adult Tobacco Survey India reveal that more than 35 per cent of total population use tobacco in various forms. Among them 21 per cent adults use only smokeless tobacco, nine per cent use smoke and five per cent use both smokeless and smoke. ^4,8^ The prevalence among male is 48% and among female is 20 per cent. ^8^

The present study aims to assess point prevalence of tobacco smoking among GATS - 1 study population and assess the association between socio-demographic factors and tobacco use in the six geographical regions of India.

## METHODS AND DATA

This is a cross sectional comparative research design, using nationally representative population based household GATS-1 data on tobacco use in India. The data on tobacco use was compiled from GATS-1 reports, which is available under public domain.

GATS 2009-10 reports provides list of regions and states (GATS report 2009-10 Table 3.5 pg.24). ^8^ Through systematic random sampling, every fourth state given in list of states from each region was chosen for analysis. In case of west region which has only three states the last one i.e. Goa was chosen.

Prevalence of tobacco smoking among study population was calculated and used for comparison between the six regions. Bi-variate analysis was used for comparative analysis among the six regions. Logistic regression was used to examine the association between socio-demographic factors (sex, education, place of residence, working status, awareness) and tobacco smoking in each zone (outcome variable).

In this manner; Chandigarh was selected to represent Northern, Madhya Pradesh (Central), Bihar (Eastern), Manipur (North-East), Goa (Western), and Tamil Nadu to represent Southern region of India (Table 1).

**Table 1.** GATS 1 sample population coverage.

| **Zone** | **State** | **N** |
| --- | --- | --- |
| North-East (NE) | Manipur | 1585 |
| Central (C) | MP | 2089 |
| Eastern (E) | Bihar | 2618 |
| Southern (S) | Tamil Nadu | 2650 |
| Northern (N) | Chandigarh | 2210 |
| Western (W) | Goa | 1931 |

## RESULTS

### Comparing prevalence in the six regions

Table 2 provides details about current use of tobacco in smoke form. From sample of total 13083 covered in all six regions, 12 per cent (1583 individuals) had reported current use tobacco in the form of smoking. Majority of tobacco smokers were from North-eastern region (Manipur; 23%) followed by Central (Madhya Pradesh: MP – 14%), Eastern (Bihar-12%), Southern (Tamil Nadu; 11%), and Northern (Chandigarh; 10%). The Western region (Goa) had lowest prevalence of tobacco smoking (5%).

**Table 2.**
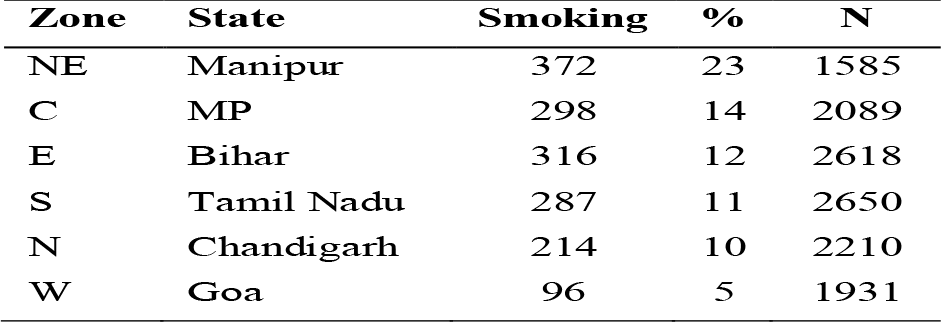
Prevalence of smoking in six regions.

| Zone | State | Smoking | % | N |
| --- | --- | --- | --- | --- |
| NE | Manipur | 372 | 23 | 1585 |
| C | MP | 298 | 14 | 2089 |
| E | Bihar | 316 | 12 | 2618 |
| S | Tamil Nadu | 287 | 11 | 2650 |
| N | Chandigarh | 214 | 10 | 2210 |
| W | Goa | 96 | 5 | 1931 |

**Table 3.** Logistic regression for Manipur (NE)

| Characteristics | Exp (B) | P-value |
| --- | --- | --- |
| **Sex** |  |  |
| Male | 0.706 | 0.000* |
| Female ® |  |  |
| **Place of residence** |  |  |
| Rural | 0.827 | 0.334 |
| Urban® |  |  |
| **Education** |  |  |
| Illiterate | 0.16 | 0.000* |
| Literate® |  |  |
| **Working status** |  |  |
| Not working | 2.418 | 0.001* |
| Working® |  |  |
| **Smoking causes stroke** |  |  |
| Yes | 1.836 | 0.012* |
| No ® |  |  |
| **Causes heart attack** |  |  |
| Yes | 1.421 | 0.35 |
| No ® |  |  |
| **Causes lung cancer** |  |  |
| Yes | 0.913 | 0.887 |
| No ® |  |  |
| **Causes serious illness** |  |  |
| Yes | 0.811 | 0.656 |
| No ® |  |  |
| **Constant** | 12.9 | 0 |
| -2 Log likelihood | 722.6 |  |

\* $p \leq 0.05$ . ®: Reference category

### Predictors of smoking in the six regions

#### North-eastern

Table 3 presents logistic regression for Manipur (North-eastern region). Results for logistic regression showed that in Manipur factors such as sex, education, working status, awareness of smoking results in stroke, had significant association with current use of tobacco in the form of smoking. Males were 1.4 times more likely to smoke tobacco than females. As compared to literate individuals; illiterates individuals 6 times more likely to smoke tobacco. Similarly, likelihood of smoking tobacco was higher in not working individuals than working individuals.

Individuals who were aware about health consequences of smoking such as stroke were less likely to smoke than those who were not aware about this causal relationship.

#### Central

For Madhya Pradesh representing central region; logistic regression showed that factors such as sex, place of residence, education awareness about smoking causes heart attack, had significant association with current tobacco smoking. It can be seen that Males had 19 times more likelihood to smoke tobacco than females. As compared to urban areas, more individuals from rural areas smoked tobacco. Illiterates individuals had two times more chances of smoking tobacco than educated individuals. Individuals who were aware about health consequences of smoking such as heart attack were less likely to smoke than those who were not aware about this causal relationship (Table 4).

**Table 4.** Logistic regression for MP (C)

| Characteristics | Exp (B) | P-value |
| --- | --- | --- |
| **Sex** |  |  |
| Male | 0.051 | 0.000* |
| Female ® |  |  |
| **Place of residence** |  |  |
| Rural | 0.630 | 0.015* |
| Urban® |  |  |
| **Education** |  |  |
| Illiterate | 0.461 | 0.000* |
| Literate® |  |  |
| **Working status** |  |  |
| Not working | 1.355 | 0.270 |
| Working® |  |  |
| **Smoking causes stroke** |  |  |
| Yes | 0.941 | 0.804 |
| No ® |  |  |
| **Causes heart attack** |  |  |
| Yes | 1.859 | 0.040* |
| No ® |  |  |
| **Causes lung cancer** |  |  |
| Yes | 1.188 | 0.639 |
| No ® |  |  |
| **Causes serious illness** |  |  |
| Yes | 1.641 | 0.438 |
| No ® |  |  |
| **Constant** | 20.941 | 0 |
| -2 Log likelihood | 804 |  |

**Table 5.** Logistic regression for Bihar (E)

| Characteristics | Exp (B) | P-value |
| --- | --- | --- |
| **Sex** |  |  |
| Male | 0.249 | 0.000* |
| Female ® |  |  |
| **Place of residence** |  |  |
| Rural | 0.550 | 0.006* |
| Urban® |  |  |
| **Education** |  |  |
| Illiterate | 0.587 | 0.011* |
| Literate® |  |  |
| **Working status** |  |  |
| Not working | 1.707 | 0.081 |
| Working® |  |  |
| **Smoking causes stroke** |  |  |
| Yes | 0.918 | 0.768 |
| No ® |  |  |
| **Causes heart attack** |  |  |
| Yes | 1.186 | 0.671 |
| No ® |  |  |
| **Causes lung cancer** |  |  |
| Yes | 1.195 | 0.776 |
| No ® |  |  |
| **Causes serious illness** |  |  |
| Yes | 2.090 | 0.199 |
| No ® |  |  |
| **Constant** | 11.4 | 0 |
| -2 Log likelihood | 769.8 |  |

\*p≤0.05. ®: Reference category

#### Eastern

In Bihar; logistic regression showed that factors such as sex, place of residence, education had significant association with current use of tobacco in the form of smoking. It can be seen that, males were four times more likely to smoke tobacco than females. Individuals from rural 1.5 time more likely to smoked tobacco than individuals from urban areas. As compared to literate; illiterate individuals were more likely to smoke tobacco (Table 5).

#### Southern

In Tamil Nadu; logistic regression showed that factors such as sex and working status had strong association with tobacco smoking. It can be seen that, males were 250 times more likely to smoke tobacco than females. Similarly, non-working individuals had more odds of smoking than those who were working (Table 6).

**Table 6.** Logistic regression for Tamil Nadu (S)

| Characteristics | Exp (B) | P-value |
| --- | --- | --- |
| **Sex** |  |  |
| Male | 0.004 | 0.000* |
| Female ® |  |  |
| **Place of residence** |  |  |
| Rural | 0.990 | 0.955 |
| Urban® |  |  |
| **Education** |  |  |
| Illiterate | 0.604 | 0.059 |
| Literate® |  |  |
| **Working status** |  |  |
| Not working | 7.829 | 0.001* |
| Working® |  |  |
| **Smoking causes stroke** |  |  |
| Yes | 0.797 | 0.441 |
| No ® |  |  |
| **Causes heart attack** |  |  |
| Yes | 0.952 | 0.930 |
| No ® |  |  |
| **Causes lung cancer** |  |  |
| Yes | 0.892 | 0.897 |
| No ® |  |  |
| **Causes serious illness** |  |  |
| Yes | 1.132 | 0.917 |
| No ® |  |  |
| **Constant** | 1026 | 0 |
| -2 Log likelihood | 848.1 |  |

\*p≤0.05. ®: Reference category

#### Western

Western region (Goa) had lowest prevalence of tobacco smoking. The logistic regression showed that factors such as sex and education had significant association with current use of tobacco in the form of smoking. In goa, males 20 times more likelihood to smoke tobacco more than females. Similarly, as compared to literate; illiterates had six times more odds of smoking tobacco (Table 7).

**Table 7.** Logistic regression for Goa (W)

| Characteristics | Exp (B) | P-value |
| --- | --- | --- |
| **Sex** |  |  |
| Male | 0.058 | 0.000* |
| Female ® |  |  |
| **Place of residence** |  |  |
| Rural | 0.980 | 0.951 |
| Urban® |  |  |
| **Education** |  |  |
| Illiterate | 0.152 | 0.000* |
| Literate® |  |  |
| **Working status** |  |  |
| Not working | 5.281 | 0.996 |
| Working® |  |  |
| **Smoking causes stroke** |  |  |
| Yes | 1.444 | 0.408 |
| No ® |  |  |
| **Causes heart attack** |  |  |
| Yes | 0.778 | 0.640 |
| No ® |  |  |
| **Causes lung cancer** |  |  |
| Yes | 0.344 | 0.310 |
| No ® |  |  |
| **Causes serious illness** |  |  |
| Yes | 2.810 | 0.354 |
| No ® |  |  |
| **Constant** | 230.9 | 0 |
| -2 Log likelihood | 299.1 |  |

\*p≤0.05. ®: Reference category

## DISCUSSION AND CONCLUSION

The study found that, North-eastern, Central and Eastern regions of India had higher prevalence of smoking. Further in all six representative states factors such as sex, place of residence, literacy status, working status had had significantly higher odds of smoking tobacco. For all states, it can be seen males smoked tobacco more than females. In some regions, place of residence had significant association with tobacco smoking. Where as compared to individuals living in rural areas were more likely to smoke than those who lived in urban areas. Similarly, as compared to literate population; illiterates had more odds of smoking tobacco. working status was also a significant predictor for tobacco smoking. Where those who were not working were more likely to smoke tobacco than working individuals. In some states, impact of health awareness about negative consequences of tobacco smoking can be seen. Particularly in North-eastern, Central and Western regions; those who were aware about causality between tobacco smoking and stroke, heart attack, and serious illness were less likely to smoke tobacco.

Secondary data analysis of GATS I reveals quite interesting facts that can be used for designing intervention that will be useful in advocating behavior change communication for cessation of tobacco smoking. Lack of awareness about negative health implications of tobacco smoking needs to be addressed with quick and effective legislative as well as community based interventions.

Trend of higher proportion in rural population in all states representing the six regions is another area which needs immediate attention. Strategies intervening demand supply chain are imperative. As high prevalence of tobacco use in rural population can be attributed to is easier access to tobacco products than urban settings. ^9,10^ Other factors that contribute to high smoking in rural areas are lower incomes and levels of educational attainment than urban population. ^11^ All these factors needs to be addressed by imposing regulatory mechanisms particularly in rural areas. Similarly higher use in males than in females reflects in higher proportion of men dying of tobacco related morbidity and mortality.

The study concludes that states in the north-east, central and eastern India region had high prevalence of tobacco smoking. Considering the socio-demographic factors, gender, place of residence, literacy, working status were the important predictors for tobacco smoking. State-specific policies are required to curb the production, use, sale, and marketing of tobacco products. ^12^ In order to curb addictive behaviors, the development and implementation of programmes to increase awareness among adults regarding the harmful use of tobacco must be developed and implemented with the help of local nongovernmental organizations (NGOs). ^12^ Further, community-based interventions to reduce second-hand smoking and the overall use of tobacco must be designed and implemented through the channels of local healthcare facilities. ^12^

## Data Availability

All data produced are available (under public domain) online at: https://ntcp.mohfw.gov.in/assets/document/surveys-reports-publications/Global-Adult-Tobacco-Survey-India-2009-2010-Report.pdf

